# The effectiveness of early diagnostic tools for neural lesions in leprosy An observational cohort study

**DOI:** 10.1101/2020.11.19.20234963

**Authors:** Luciano Henrique Vieira Cegana, Susilene Maria Tonelli Nardi, Luciana Pascoeto, Vania Del Arco Paschoal

## Abstract

Cegana LHV, Nardi SMT, Pascoeto L, Paschoal VDA. The effectiveness of early diagnostic tools for neural lesions in leprosy. 37p. MANUSCRIPT, 2020. https://doi.org/10.1101/2020.11.19.20234963

**Introduction:** Leprosy can cause different lesions in peripheral nerves and inervatory structures.

**Objectives:** To analyse the effectiveness of evaluation protocols used to identify neural lesions in leprosy such as Degree of Physical Disability (DPD), Simplified Neurological Assessment (SNA) and propose to use Neurodynamic Assessment (NDA).

**Method:** Descriptive analytical study, associative, with 27 individuals treated in two outpatient leprosy clinics in São Paulo State, between 2017 and 2019, and 27 individuals from the paired control group. The Mann-Whitney, Multivariate Linear Regression and association between variables and P<0.05 values were used.

**Results:** The test that most captured the neurological alterations was the SNA, with 22 (81.5%) in the upper limbs (ULs) and 25 (92.6%) in the lower limbs (LLs), followed by the NDA, with 20 (74.1%) in the ULs and 11 (40.7%) in the LLs. The DPD showed handicap in the hands of 16 (59.2%) individuals and in the feet of 17 (62.9%) individuals, and they have expressed sensitivity. DPD showed agreement with SNA in 21 (77.8%) of the cases in ULs (p=0.010) and 19 (70,4%) of the cases in LLs (p=0.060). DPD and NDA showed that in 19 (70.4%) of the patients evaluated there was concordance of results in ULs (p=0.143); 9 (33.3%) in LLs (p=0.125). SNA and the NDA in the ULs found agreement in 21 (77.8%); 11 (40.7%) (p=0.786) in the LLs.

**Conclusion:** The three assessment instruments are specific and will hardly produce false positive tests. DPD can produce more false negatives than SNA. If there is an instrument to be chosen, it should be the SNA, since it is more sensitive, more accurate and has a less negative likelihood ratio. Neurodynamic tests were positive in 7.4% of individuals while there were still no changes in the SNA; afterwards, these changes appeared.

**Descriptors:** Neural mobilization; Leprosy; Peripheral nerves; Disability; Pain; Physical therapy.

## INTRODUCTION

Among infectious diseases, leprosy is considered one of the main causes of physical disabilities, due to its potential to cause neural injuries^[1]^.

Worldwide, 208,619 new cases of the disease were reported to the World Health Organization (WHO) in 2018, 30,957 occurred in the Americas region and 28,660 (92.6% of the total from the Americas) were reported in Brazil. New cases, which were assessed for its Degree of Physical Disability (DPD), totaled 86.5% (n=24,780); 2,109 (8.5%) had visible physical deformities (DPD 2)^[2]^.

The lack of new diagnostic tools and new drugs, limited knowledge about strategic areas of transmission and unsatisfactory tools for managing the complications caused by leprosy have been an obstacle to its control. Therefore, more coordinated research efforts are still needed^[3]^.

The diagnosis of leprosy is based on the presence of at least one of three cardinal signs: a) lesion(s) and/or area(s) of the skin with changes in thermal and/or pain and/or tactile sensitivity; or b) thickening of the nerve trunks of the peripheral nerves in the upper and lower limbs, associated with sensitivity and/or motor changes in extremities, with decreased muscle strength in the myotomes supplied by these and/or autonomic nerves; or c) presence of *Mycobacterium leprae* bacilli, confirmed by intradermal smear microscopy or skin biopsy^[4]^.

Sometimes neurological involvement is preponderant when compared to a dermatological condition not very significant^[5]^. The most affected nerves are the ulnar in the elbow and the fibular in the head of the fibula, followed by the ulnar sensory, superficial and sural fibular branches^[6]^.

In daily practice, a greater occurrence of sensory changes over motor changes is found, as well as a slight presence of deformities^[7]^. The sensory and motor actions of *M*.*leprae* in the upper and lower limbs cause secondary injuries such as fingers ^[8]^ and toes in flexion and “fallen hands and foot”^[9]^.

Although the prevalence of leprosy is decreasing, cases that no longer have the active disease, but which present neurological sequelae, continue to be monitored in health services^[10]^. Leprosy reactions are one of the main causes of the onset of disabilities^[11]^. It is essential to assess the integrity of neural function at the time of diagnosis, in the occurrence of reactive states, at discharge due to cure (end of multidrug therapy) and during 5 years after discharge^[7]^.

Peripheral neuropathies in leprosy are a triggering factor for physical disabilities, and some evaluation protocols can be used for their prevention, such as the Degree of Physical Disability of the World Health Organization (DPD) and the Simplified Neurological Assessment (SNA) of the Brazilian Ministry of Health^[12]^. The routine protocols recommended by the Brazilian Ministry of Health are subjective and, in view of that, other tests could complement the diagnostic examination of peripheral neuropathy such as the Neurodynamic Assessment (NDA)^[7]^.

The DPD is an epidemiological indicator, not very sensitive, but it can define the severity of neurological changes. The SNA assesses muscular strength and sensitivity of eyes, hands and feet, in addition to the palpation of the possibly affected nerves and the morphological evaluation of the limbs^[7]^.

The Neurodynamic Assessment (NDA) is used for injury diagnosis and treatment of the peripheral nervous system and the structures innervated by it^[13]^; the interface between the musculoskeletal system and the peripheral nervous system is used, so the movements applied to the musculoskeletal system mobilize the structures of the peripheral nervous system^[14]^.

The use of neurodynamic tests has helped in the diagnosis of neural lesions to identify the origin of the lesions, but little is known about the results of the neurodynamic tests applied to patients with leprosy. Thus, we suggest using the mobilization of the nervous system in clinical practice as a tool to assess peripheral nerves that may be affected by leprosy.

### OBJECTIVES

Analyzing the effectiveness of the evaluation protocols used to identify neural lesions in leprosy, such as the Degree of Physical Disability (DPD) recommended by the World Health Organization, the Simplified Neurological Assessment (SNA) and propose to use the Neurodynamic Assessment (NDA)

Describing the sociodemographic and clinical variables of the groups studied and check whether there is a statistical association between the results of the evaluation tools: the Degree of Physical Disability recommended by the World Health Organization (DPD), the Simplified Neurological Assessment (SNA) and the positive Neurodynamic Assessment (NDA) in individuals who have or had leprosy, in addition to determining the specificity and sensitivity of each of the assessment protocols.

## METHODS

Associative, analytical, descriptive research, following case-control design, approved by the FAMERP Ethics and Research Committee in accordance with the requirements of National Resolution No. 196/96, Opinion No. 2,469,355. Participants were asked to sign the Informed Consent Form (ICF) in accordance with Ordinance No. 466/2012.

The study included all leprosy patients diagnosed between 2017 and 2019, that agreed to sign ICF, N=27 of both sexes and different ages, in one medium (50 to 100 thousand inhabitants) and one large (100,001 to 900,000 inhabitants) Brazilian municipality (Votuporanga and São José do Rio Preto), under treatment or undergoing chemotherapy. A control group of 27 people, chosen from the population, without a diagnosis of leprosy, was evaluated pairing age and sex, thus totaling 54 people. Data collection went from December 2017 to November 2019, until three evaluations were completed, with an average interval of 3 months between evaluations.

We selected as exclusion criterion for both groups those with compressive syndromes of the central nervous system (CNS) and the peripheral nervous system, other diseases of the CNS such as stroke or degenerative syndromes, diabetics and alcoholics with altered sensitivity and people/patients who refused to sign the informed consent form.

Initially, a patient profile data sheet, extracted from their medical records, was used, with the name, address, age, sex, date of the beginning of treatment, date of discharge, clinical classification of leprosy and type of treatment. In the control group, name, address, age, sex and existing morbidities were used. Evaluation instruments were applied:

- **Simplified Neurological Assessment (SNA)** is a protocol recommended by the Brazilian Ministry of Health, and it contemplates dermatoneurological exploration, the evaluation of eyes, hands and feet sensitivity, and the evaluation of the motor function^[15]^. In this clinical examination, the integrity of the skin and its nutrition are assessed. Testing quick perception of a light touch and/or deep pressure^[15]^. It is recommended to use the Semmes-Weinstein monofilament set (6 monofilaments: 0.05g, 0.2g, 2g, 4g, 10g and 300g) in the sensitivity assessment points in hands (palmar surface and back of the hand) and feet (plantar surface and instep near the hallux and the second toe)^[15]^. To assess motor strength, it is recommended to perform manual testing of muscle strength in the muscle tendon unit during movement and the ability to oppose gravity and manual resistance in each muscle group from a specific nerve myotome. Ulnar nerve (Abductor digiti minimi), median nerve (abductor pollicis brevis), radial nerve (wrist extensors), fibular nerve (tibialis anterior, extensor digitorum longus, extensor hallucis longus, fibularis tertios).

The grading criteria for muscle strength can be expressed from 0 to 5 following the Kendall scale for the degree of muscle strength^[15]^. (Appendix 1)

- **WHO Degree of Physical Disability (DPD**) indicates the loss of protective sensitivity, muscle strength and / or visible deformities in the face, upper and lower limbs, varies between Degree 0 (no leprosy-related physical disability), I (decreased strength and/or loss of sensation) or II (presence of visible disabilities and deformities) according to the severity of sensory and/or motor and/or morphological changes caused by neural lesions of leprosy^[15]^.
- **Neurodynamic tests**: straight leg raises (SLR), Slump Test and upper limb tension tests (ULTT) 1, 2b and 3^[16]^.

1. The straight-leg raise test (SLR) is performed with the patient in the supine position, with the trunk and hips in neutral positions. The examiner places one hand under the Achilles tendon and the other above the knee. The hip is flexed with the knee held in extension until it reveals a predetermined symptomatic response, or until it reaches its hip range of motion (ROM) limit. The ROM value should be compared to the SLR of the contralateral limb, and to what is considered as normal. If pain, especially lower back pain, is reported during the test, the most likely cause is a herniated disc or a central pathology causing compression. The normal value of SLR in normal individuals varies from 50° to 120°^[16]^. But if pain is reported in the limb following the nerve path, we consider the test positive in individuals with leprosy, as neural tension can be caused by compression or incarceration of the peripheral nerve. Painful expression or reproduction of symptoms usually in the course of innervation. Do not confuse with the discomfort of hamstring stretching, because when maximal knee extension increases posterior thigh pain, it did not increase muscle tension, measured indirectly using EMG. The test is considered positive when the patient reports any discomfort/pain (pins and needles and sensation changes, pulling, tension, pain, numbness or tingling, that radiates inferiorly through the posterior/lateral sides of the leg, often associated with paresthesia) or when the physical therapist encounters resistance to movement ^[17]^.
2. The Slump Test should be performed with the patient seated, with the thighs fully supported, the knees together and the hands together on the back. The patient is asked to flex the thoracic and lumbar spine, keeping the sacrum in a vertical position and, soon after, also perform cervical flexion. The physiotherapist puts pressure on the cervical region in order to accentuate flexion. Then, the patient performs an active knee extension, associated with dorsal flexion of the ankle. Cervical flexion is slowly released, and the painful response must be carefully evaluated. The symptoms must be noted at each stage and must also be performed for the other member. ROM and painful responses should be assessed^[16]^.
3. The upper limb tension test (ULTT1) (median neural tension) is described as a brachial plexus tension test or a median nerve test. It is performed with the patient in the supine position. The examiner exerts force to depress the scapular waist, which has an external rotation and 110° abduction of the glenohumeral articulation, elbow extension, radioulnar supination and wrist and fingers extension, the inclination of the cervical to the opposite side was suppressed in our assessment to isolate core involvements. ULTT1 tests are considered positive if present: complaint at deep elongation or pain in the cubital fossa that extends down the anterior and radial part of the forearm and to the radial side of the hand; tingling sensation in the first four fingers; stretching in the anterior shoulder area^[16]^.
4. The upper limb tension tests (ULTT 2b) assess the radial nerve with the patient in the supine position. The examiner holds the elbow and wrist of the patient. Using the thigh, the examiner depresses the scapular waist and internally rotates the shoulder, extends the elbow, flexes the wrist, fingers and thumb^[16]^.
5. The upper limb tension test (ULTT 3) (neural tension of the ulnar) is performed with the patient in the supine position, keeping the wrist of the patient extended and the forearm supine, performing an elbow flexion. After positioning the patient, the therapist performs a shoulder depression associated with an external rotation. Shoulder abduction is added (so that the hand is close to the ear of the patient). In our assessment, we suppressed lateral flexion of the cervical to isolate central pathologies. The test is considered positive when the patient reports any discomfort/pain or when the physical therapist encounters resistance to movement. Pain is intermittent, deep and burning in quality. Also, a cold, tingling feeling extended distally from the medial elbow to the little finger ^[18]^. When the patient has any of this reported symptoms, the test is completed and the physiotherapist keeps the limb of the patient positioned in the range in which pain or resistance to movement is reported to perform a goniometer measurement^[14]^.

The leprosy group underwent 3 evaluations, quarterly and the control group was evaluated in a single moment. None of the subjects were in reaction episode at the time of the evaluation. The application of the data collection instrument took 40 to 50 minutes and was performed by a single examiner.

### Data analysis

- The Simplified Neurological Assessment was assigned with a value “without changes” or with the value “with changes” for the upper and lower limbs. We considered that the change in sensitivity in hands decreased when the individual didn’t feel the 0,02g green monofilament. The decrease in plantar sensitivity was considered when didn’t feel the 0,5g blue monofilament. The muscle strength domain was also considered to perform SNA grouping and was coded as “altered” when muscle strength was less than 5, according to the Kendall scale, identifying a muscular paresis and therefore a neurological alteration ^[15]^. The palpation was marked as altered when there was pain, thickening, or both.
- We synthesize the results of the Neurodynamic Assessment in positive and negative tests, considering the set of tests for the upper and lower limbs one apart to the other.
- The degree of physical disability of the WHO was classified as “*without disability*” when it was Degree 0, and was grouped as “*disabled*” when its results were Degrees 1 or 2.

After tabulation of the data collected in this work, 2 analytical and statistical functions were performed: descriptive and inferential. In a descriptive way, the profile of the studied sample was drawn, considering the analyzed variables and their consequences, and the data was replicated in an absolute and relative way.

In the inferential scope, the analysis of independence and prediction between variables was drawn as a statistical objective. In addition, the Mann-Whitney U test and Multivariate Linear Regression were used with the results of independence between the proposed variables, taking into account the P-values (significance P≤ 0.05). All analyses were obtained using the SPSS Statistics Software (Version 23) and were linked to the features of the Excel tool (version 2016).

Statistical analysis and synthesis of the results were performed on a Venn-Euler diagram. To perform the descriptive analysis and verify the association between SNA, DPD and NDA results, EPI INFO 7.1 statistical software was used.

To validate the diagnostic tests, calculations of sensitivity, specificity, positive and negative likelihood ratios were performed using the Virtual Health Library (*Biblioteca Virtual de Saúde*, BVS) virtual calculator^[19]^. Sensitivity, specificity and accuracy vary from 0-1. A sensitive and specific test is one which is positive in the presence of pathology and negative in the absence of pathology^[18,20]^.

Likelihood ratios show the performance of a diagnostic test. The Positive Likelihood Ratio (PLR) varies from 1 to infinity, the higher your result, the better the test. The Negative Likelihood Ratio (NLR) varies from 1 to 0, the closer to 0, the better the test^[21]^.

## RESULTS

In this study, all 27 individuals diagnosed with leprosy who were notified in the years 2017 to 2019 participated (8 in the municipality of Votuporanga, 19 in the municipality of São José do Rio Preto), in addition to 27 volunteers matched for age and sex in the control group, totaling 54 people. Among the intervention group, 17 (63%) were male. The ages varied between 23 and 88 years in the control group and between 23 and 87 years in the intervention group, with a mean age of 53.1 (SD 17.6). The age median in the control group was 57 while in the intervention group it was 55.

The age distribution was identical in the control and intervention groups due to the pairing (n=54) (p-value 0.945). There was no difference in sex (p-value 1.00). In the intervention group there were 19 (70.4%) active workers, in the control group it was 22 (81.5%) (p-value 0.344).

Regarding leprosy, 20 (74.1%) of the participants in the intervention group had the dimorphous and Virchow’s leprosy clinical forms, with multibacillary multidrug therapy (74.1%). Seven (25.9%) patients had a reaction phenomenon to leprosy during the treatment, but not at the time of the evaluations. 25 (92,6%) still remained under drug treatment, 11 (40,7%) were evaluated in less than 30 days after diagnosis, 4 (14,8%) were evaluated in less than 60 days after diagnosis, 1 (3,7%) were evaluated in less than 90 days after diagnosis, 3 (11,1%) before 180 days after diagnosis, 3 (11,1%) before 270 days after diagnosis and 3 (11,1%) before completing 365 days between diagnosis and first evaluation. 2 (7,4%) patients were evaluated after de discharge.

The participants in the intervention group showed neurological changes, at least in one site of the upper limbs, assessed by the three types of exams. The test that most captured data was the SNA with 22 (81.5%) participants, followed by the Neurodynamic Assessment, where 20 (74.1%) were altered, and the DPD showed deficiency in the hands of 16 (59.2%) individuals.

As assessing the lower limbs, it appears that in the intervention group the participants showed neurological changes in the three assessments studied. Neurological changes were more perceived by SNA, attested in 25 (92.6%) participants. The Degree of Physical Disability showed a disability in 17 (62.9%) individuals. In the Neurodynamic Assessment, 11 (40.7%) showed alterations. In the control group, no neurological changes were identified by the 3 instruments, confirming the reliability of the sample (Table 1).

**Table 1.** Distribution of the results of the Simplified Neurological Assessment, degree of hand and foot disabilities, neurodynamic test of the upper and lower limbs in the intervention group. (N=27)

| TESTS | RESULTS | LIMBS |  |  |  |  |  |  |  |
| --- | --- | --- | --- | --- | --- | --- | --- | --- | --- |
|  |  | Intervention Group |  |  |  | Control Group |  |  |  |
|  |  | Upper |  | Lower |  | Upper |  | Lower |  |
|  |  | n | % | n | % | n | % | n | % |
| **Simplified Neurological Assessment** | No alteration | 5 | 18.52 | 2 | 7.41 | 27 | 100 | 27 | 100 |
|  | Alteration | 22 | 81.48 | 25 | 92.59 | 0 | 0 | 0 | 0 |
| **Degree of Physical Disability** | No disability (Degree zero) | 11 | 40.74 | 10 | 37.04 | 27 | 100 | 27 | 100 |
|  | Disability (Degree I and II) | 16 | 59.26 | 17 | 62.96 | 0 | 0 | 0 | 0 |
| **Neurodynamic Assessment** | Negative | 7 | 25.93 | 16 | 59.26 | 27 | 100 | 27 | 100 |
|  | Positive | 20 | 74.07 | 11 | 40.74 | 0 | 0 | 0 | 0 |

In Figure 1, using the Venn-Euler diagram, it can be seen that 51.8% (14 of 27) of patients were diagnosed with neural problems in the upper limbs using the three analysis instruments. When we use only two associated assessments, the number of cases with changes decreases. We also observed that the instruments individually captured different people with problems, 2 (7.4%) with SNA and 2 with NDA; that is, 2 individuals that only had their neural lesions identified because they passed through SNA and another 2 different individuals only had neural injury identified because they were subjected to NDA.

**Figure 1.**
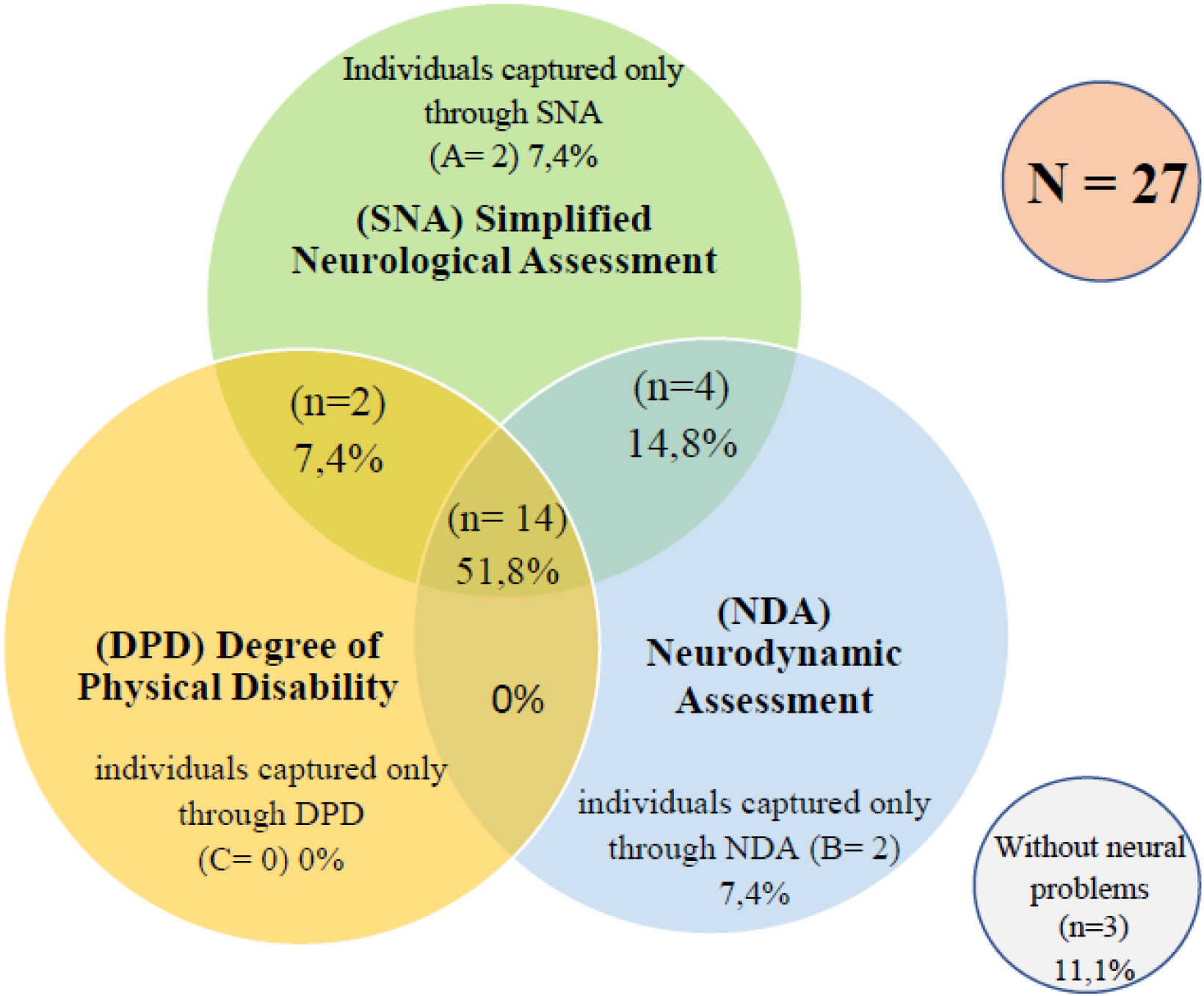
Intersection of three assessments used to check upper limbs (ULs) neurological injuries of individuals with leprosy using the Venn-Euler Diagram. **Caption**. SNA: Simplified Neurological Assessment; DPD: Degree of Physical Disability; NDA: Neurodynamic Assessment. SNA identified neural injury in 22 individuals, NDA in 20 individuals and DPD in 16. The intersection zones correspond to individuals captured by 2 or 3 assessment instruments. The numbers outside the intersections mean the individuals who are captured only by that specific assessment. A = individuals captured only by SNA, B = individuals captured only by NDA, C = individuals captured only by DPD.

In Figure 2, using the Venn-Euler diagram, it can be seen that 18.5% (5 of 27) of patients were diagnosed with neural problems in the lower limbs using the three analysis instruments. When using two instruments, SNA and NDA, the number of identified cases remains with 5, (18.5%); using NDA and DPD it is 0 (0%), as they do not identify any individual who is also not identified by SNA. Using the DPD instrument and the SNA together identifies more individuals with neural lesions 12 (44.4%). We also observed that the instruments individually captured different people with problems, 3 (11.1%) with SNA and 1 (3.7%) with NDA; that is, three individuals only had their neural lesions identified because they passed through SNA and the neural lesion of another different individual was only identified because he was subjected to NDA.

**Figure 2.**
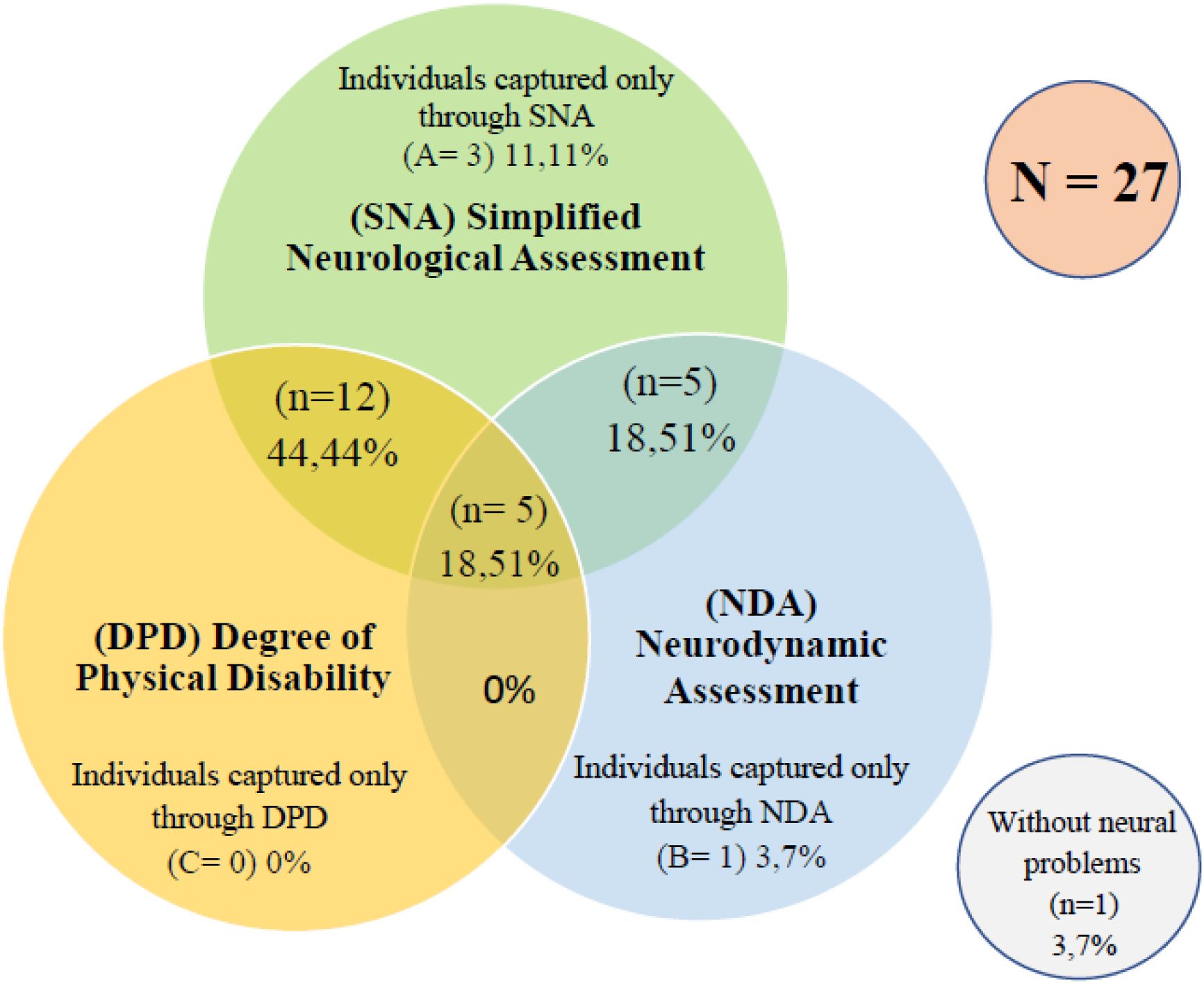
Intersection of the results obtained from the three assessments used to verify neurological injuries in the lower limbs (LLs) of individuals with leprosy **Caption**. SNA: Simplified Neurological Assessment; DPD: Degree of Physical Disability; NDA: Neurodynamic Assessment. SNA identified neural injury in 25 individuals, NDA in 11 individuals and DPD in 17. The intersection zones correspond to individuals captured by 2 or 3 assessment instruments. The numbers outside the intersections mean the individuals who are captured only by that specific assessment. A= individuals captured only through SNA, B = individuals captured only through NDA, C = individuals captured only through DPD

### Sensitivity, specificity, positive and negative likelihood ratio

To define the characteristics of the tests as a diagnosis, we used the concepts of sensitivity, specificity and positive and negative likelihood ratios considering the ULs and the LLs separately, based on the distribution of the test results, in order to provide evidence on which clinical practices can be based. (Table 2)

**Table 2.** Measures of sensitivity, specificity, accuracy, positive and negative likelihood ratios for Simplified Neurological Assessment, Neurodynamic Assessment (NDA) and Degree of Physical Disability (DPD) in the upper and lower limbs

| Measures | ULs | SNA Test | NDA Test | DPD Test |
| --- | --- | --- | --- | --- |
| 0-1 | Sensitivity | 0.81 | 0.74 | 0.59 |
|  | Specificity | 1 | 1 | 1 |
|  | Accuracy | 0.90 | 0.87 | 0.80 |
| 1 to 0 | NLR | 0.18 | 0.26 | 0.41 |
| 1 to infinity | PLR | Infinite | Infinite | Infinite |
| Measures | LLs | SNA Test | NDA Test | DPD Test |
| 0-1 | Sensitivity | 0.92 | 0.41 | 0.63 |
|  | Specificity | 1 | 1 | 1 |
|  | Accuracy | 0.96 | 0.70 | 0.81 |
| 1 to 0 | NLR | 0.07 | 0.59 | 0.37 |
| 1 to infinity | PLR | Infinite | Infinite | Infinite |

**Caption:** SNA: Simplified neurological assessment; NDA: Neurodynamic Assessment; DPD: Degree of Physical Disability; ULs: Upper limbs; LLs: Lower limbs NLR: negative likelihood ratio; PLR: positive likelihood ratio<sup>43</sup>

Our sample tells us that the instruments used are specific because the 3 assessments obtained a value equal to 1 in both the upper limbs and lower limbs, and that they hardly produce false positives, since the positive likelihood ratio tended to infinity in the 3 assessment instruments^[21]^.

Considering the sensitivity, we saw that SNA is the most sensitive test to assess the upper limbs in individuals with leprosy, as it had a value of 0.81, followed by the NDA with 0.74. The DPD is considered least sensitive, with a value that was equal to 0.59^[20]^. In LLs, SNA remains as the most sensible over other assessment instruments, with 0.92. NDA was not a tool as sensitive for LLs as it was for ULs because we obtained a sensitivity value equal to 0.41. DPD obtained a better sensitivity, with 0.63^[20]^.

The best accuracy in ULs was obtained by SNA (0.90), followed by NDA (with 0.87) and DPD (with 0.80), whereas in LLs, the accuracy of SNA was 0.96, followed by DPD, with 0.81, and NDA, with 0.70^[20]^.

According to the Negative Likelihood Ratio (NLR) in ULs, SNA obtained the best value among the evaluations, with 0.18, while the value in the NDA and DPD was 0.26 and 0.40, respectively, showing that false negatives can occur more frequently in the NDA, and could be still larger in the DPD. In LLs, the NLR in the SNA was 0.07, meaning that it is difficult for the SNA to produce a false negative; however, the NDA can produce false negatives because the NLR was equal to 0.60; the DPD also could report no deficiency while in fact there is already a neural alteration because the NLR was equal to 0.37^[21]^.

### Degree of hand and foot disabilities compared to the Simplified Neurological Assessment and Neurodynamic Assessment of upper and lower limbs

The WHO Disability Degree indicator of the hands shows agreement in 21 (77.8%) of the cases that were assessed by SNA (P-value = 0.010). It should be noted that the SNA made the diagnosis of neural injury in 6 (22.3%) cases that, according to the DPD, did not have physical disability (degree zero of physical disability).

Regarding the association of the degree of hand impairment with the Neurodynamic Assessment in 51.8% (n=14) of the patients evaluated there were agreed positive results; in 18.5% (n=5) there were agreed negative results. The results differed in eight participants, with the Neurodynamic Assessment more frequently capturing neural changes (n=6/22.2%) than the Degree of Physical Disability (n=2/7.4%). The association was not significant (p-value=0.143).

The comparison between the degree of disability of the feet and the SNA of the lower limbs was acceptable in 62.9% (n=17) of the cases but diverged in 29.6% (n=8) cases, exposing a greater sensitivity of the SNA in showing the neural damage, despite the association not showing statistical significance (p-value=0.060).

Verifying the degree of disability of the feet and the Neurodynamic Assessment of the lower limbs showed agreement of positive results in 18.5% (n=5) of the patients evaluated and in 14.8% (n=4) of the negative results. In eighteen participants, the results differed; the Degree of Physical Disability of the feet more frequently captured neural changes (n=12/44.4%) than the Neurodynamic Assessment (n=6/22.2%). The association was not significant (p-value=0.125). (Table 3)

**Table 3.**
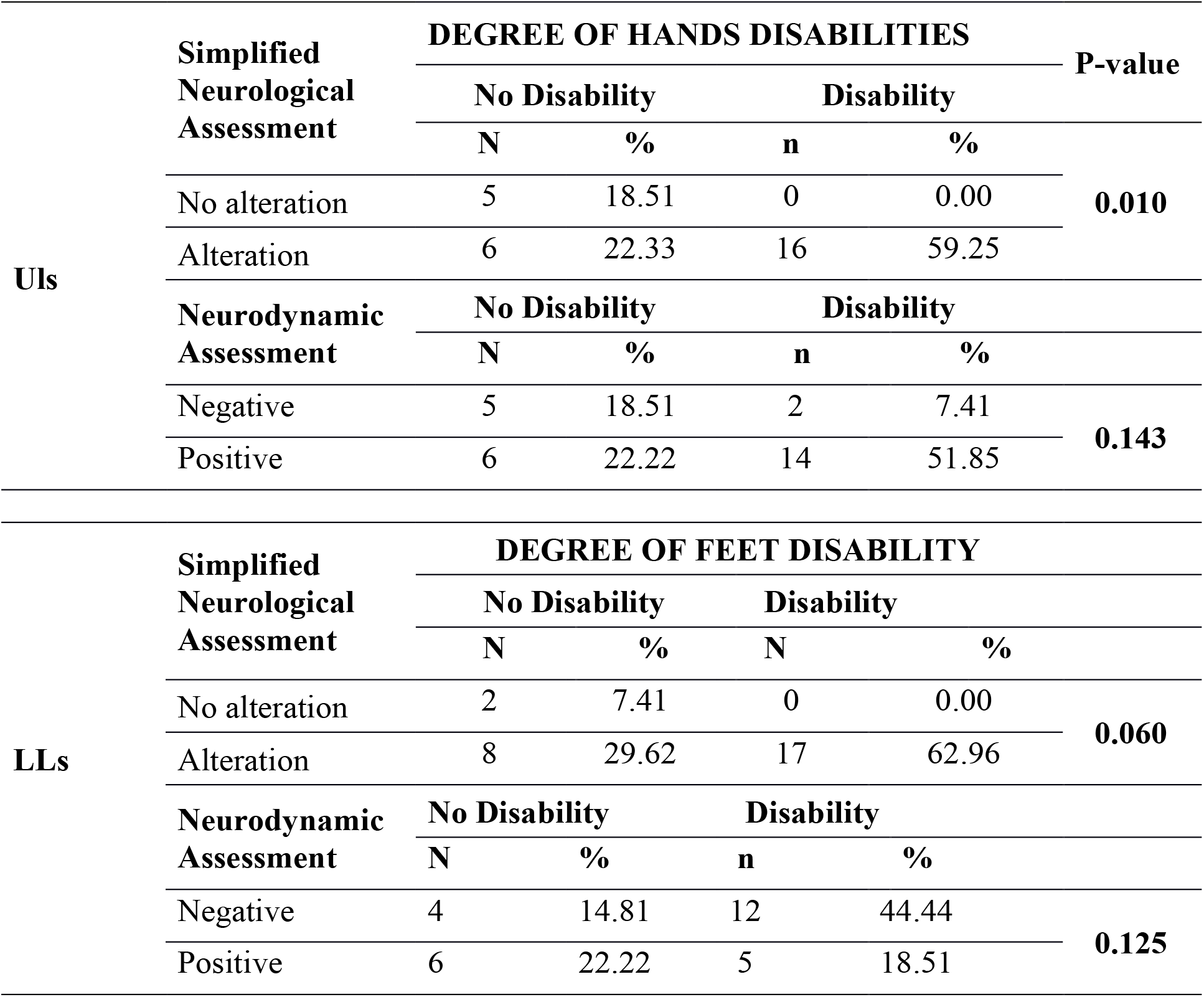
Degree of hand and foot disabilities compared to Simplified Neurological Assessment and Neurodynamic Assessment in upper limbs and lower limbs when applied to the intervention group. (n=27)

### Simplified Neurological Assessment compared to Neurodynamic Assessment in the Upper and Lower Limbs

By checking the Simplified Neurological Assessment and the Neurodynamic Assessment in the ULs, we found agreement on the positive results in 66.7% (n=18) of the cases with neurological changes and in 11.1% (n=3) of the cases without changes with negative tests. There was disagreement in six individuals. The SNA was more effective in verifying the neurological changes of the upper limbs in 14.8% (n=4) cases while the Neurodynamic Assessment identified it in 7.4% (n=2) cases.

The association between Simplified Neurological Assessment and the Neurodynamic Assessment of lower limbs showed agreement on positive results in 37% (n=10) of the patients evaluated and in 3.7% (n=1) of the negative results. In sixteen participants, the results differed, the SNA of LLs more frequently captured neural changes (n=15/55.5%) than the Neurodynamic Assessment of the lower limbs (n=1/3.7%). The association was not significant (p-value=0.786). (Table 4)

**Table 4.**
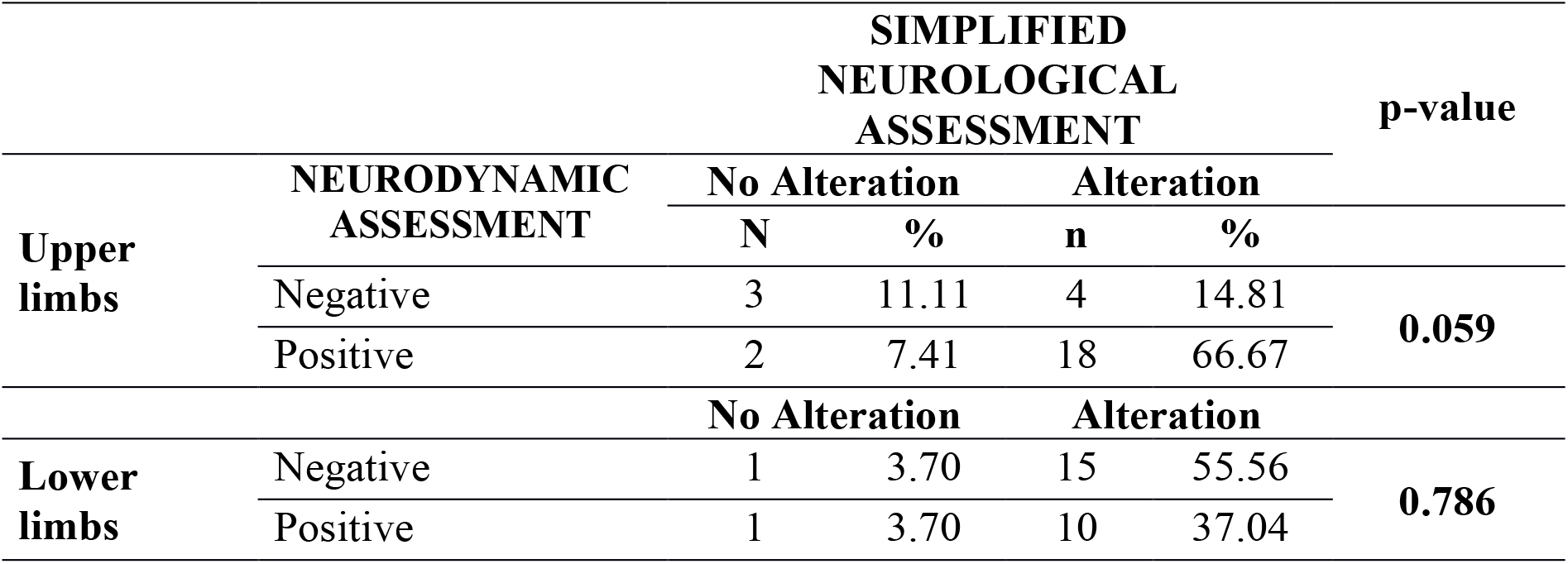
Simplified Neurological Assessment compared to Neurodynamic Assessment, in the upper and lower limbs, when applied to the intervention group.

## DISCUSSION

With this study we hoped to identify an evaluation protocol that used only human resources for its execution, facilitating the identification of neural lesions in leprosy combined with the daily practice of physical therapists who are not involved in the management of leprosy, but routinely use Neurodynamic Assessment. In this way, more suspicions and diagnoses of the disease could be made and would help in cases that present themselves as purely neural. Concerning leprosy, it is important that not only the physician suspects the disease, but any health professional.

There was no statistical difference in relation to sex, age and occupation in the two groups studied, since the pairing was performed to avoid the effect of age and sex, two recognized confounding factors for the results^[22,23]^, so a comparison was made between groups, showing similarity and equal care between them^[24,25]^.

In the intervention group, men, active, with an average age of 53.09 years and multibacillary disease were more frequent, as happened in Santarém, where they had ages above 51 years old^[26]^, being one of the sociodemographic factors that were associated with the installation of disabilities^[27]^.

Regarding neurological tests, the one that most captured neural alterations was the SNA in both upper limbs and lower limbs, highlighting the importance of the process of identifying the degree of disabilities through SNA, at the beginning of treatment and after discharge^[28]^. Studies show worsening of physical disabilities during treatment, mainly in the multibacillary form, with the lower limbs being the segments that show the most significant degree of disability evolution, justifying the imperative need for a more careful monitoring of these cases through routine assessments and interventions^[29]^.

In the univariate and multivariate analysis, there is agreement of the result found in the DPD with the SNA in the upper limbs, but the SNA detects neural lesions more sensitively. Since disability is only considered when sensitivity is decreased beyond purple monofilament and/or muscle strength is less than 5, DPD does not detect the subtle changes that suggest neural distress, and we emphasize that the SNA made the diagnosis of neural injury in 6 (22.3%) of the cases while, according to the DPD, those cases did not yet present disabilities, showing the importance of using and applying SNA.

The degree of physical disability was an innovative measure to gradually quantify physical disability in an index and were not designed to be sensitive to early changes, but with the increasingly early diagnosis of neural damage, it was necessary to improve the assessment instruments. Due to the change in the profile of patients today, these instruments need to be more sensitive to subtle changes that neurological changes show us over the course of the disease, so SNA is more sensitive in detecting neurological changes and their influences on socioeconomic and emotional aspects^[30]^.

The DPD is recommended at the beginning of treatment and at discharge, but when the patient shows a slight improvement, there is no variation in the degree of disability and the services are unable to assess whether the actions developed are actually being effective^[31]^. It must be considered that when the case is carefully evaluated and the results of the exams recorded in the SNA protocol, the diagnosis of deficiencies in the eyes, hands and feet is more accurate than when the deficiency is identified using the framework “WHO degree of physical disability”^[28]^.

As an indicator, there is a fragile and subjective comparison of the DPD with the limitation of activities, and the social participation of the patients is not included in its results. An adaptation of the indicator is necessary to develop a more current classification, based on a more comprehensive concept of disability, as is done by the International Classification of Functioning (ICF)^[32]^.

Nerve damage is associated with physical disabilities. Thus, regular monitoring of nerve function, through SNA, combined with adequate clinical management of neuritis, neuropathies and leprosy reactions, are effective strategies to prevent it^[33]^.

In the univariate analysis, as well as in the multivariate, of ULs and LLs, DPD and NDA, despite their results being relative to neural injury concordance in some cases, were not statistically significant. The NDA seems to identify 75% more injuries in the upper limbs than the DPD, and DPD seems to identify 50% more injuries in the lower limbs than the NDA.

Considering leprosy and NDA, a study of ULs found a positive NDA, mainly affecting those with a degree of physical disability level 2. Of these, they presented a decrease in the ROM of elbow flexion in the ulnar neural tension test (ULTT3) on both sides when compared to control group^[14]^.

Leprosy patients test positive when submitted to Neurodynamic Assessment of lower limbs^[25]^. But, even when the tests do not reproduce the symptoms in the affected nerves, characterizing negative tests, studies suggest that a neuropathy cannot be ruled out yet, this can mean a more severe lesion with demyelination of the fibres^[35]^ (as occurred in degree 2 patients that tested negative in the lower limbs in our analysis), producing a possible false negative result^[20]^, thus explaining the low sensitivity of the NDA in lower limbs, because low sensitivity produces more false negative results^[20]^.

Both in the univariate and multivariate analysis of ULs and LLs, when comparing the SNA with the NDA, we note that the SNA identifies the neural lesion earlier than the NDA.

According to the evidence available in 2019, it is suggested that neurodynamic tests should not be used in isolation, as a single test, to diagnose neural distress of the median nerve. They should be interpreted in the context of a loss of function tests of small fibres in a domain^[34]^. Combining anamnesis and clinical history is an important tool to make the differential diagnosis, where the combination of negative neurodynamic test results could be used to rule out a disorder in the peripheral nerves; with limited evidence, they are relevant in cervical radiculopathies^[35]^.

The need for adaptation of the peripheral nervous system (PNS) in the face of mechanical stress, stretching, slips, changes in its diameter and compressions is notorious. If this dynamic protection mechanism fails, the PNS is vulnerable to edema, ischemia, fibrosis and hypoxia, which are the causes of neural changes^[36]^.

In this sense, the scientific community highlights the need for more possibilities for neural injury investigations in leprosy, in order to diagnose and monitor the neurological changes caused by it. A study in Nigeria points out that 50% of patients who complete treatment already had neurological changes before diagnosis, according to the EHF score, but 90%, when receiving assistance and monitoring of injuries, end treatment with less disability^[37]^. Neural damage needs to be identified early and current leprosy control efforts must be intensified to ensure immediate treatment in order to reduce the burden of the disease, including deficiencies in individuals and the community. When using EHF, greater sensitivity is offered in the evaluation compared to DPD^[31]^. Physical disabilities were diagnosed in 41% of the individuals. The EHF score showed overlapping of impairments in the examined segments and proved to be more appropriate than the DPD classification system to describe the extent of the physical disabilities in the patients^[38]^.

In our study, neurodynamic tests were positive in 2 (7.4%) individuals while there were still no changes in SNA and later these changes appeared, which makes us think of the association of assessments as a way to complement the diagnosis and monitoring of neural changes. This would make professionals, when applying NDA, be able to suspect neural injury caused by leprosy and minimally perform other evaluations, making a more accurate investigation or even referring it to a specialist in the area.

Therefore, a need to create an assessment (validated or not) is necessary so that we can, through an indicator, reveal the neural lesion more precociously, given that 95% of patients have neurological changes, with musculoskeletal symptoms, which interfere in their functional capacities, causing difficulties in performing their activities of daily living and work when compared to those who have no symptoms. However, the presence of disability did not prevent or limit them from performing these activities. Even with pain, paresthesia, decreased strength and other injuries, they still perform their activities^[39]^.

## Conclusions

We conclude that the 3 assessment instruments are specific and that these assessments can and should be used in combination to expand the monitoring of neural lesions in leprosy, as there are changes that are not perceptible with one instrument but can be confirmed by another.

The Simplified Neurological Assessment (SNA) is the most sensitive instrument, with greater accuracy and it is unlikely to produce a false negative both in the assessments of the upper limbs and lower limbs. The Neurodynamic Assessment (NDA) is the second most sensitive instrument for assessing ULs, followed by the Degree of Physical Disability (DPD). In LLs, the second most sensitive instrument is DPD, followed by NDA.

Both NDA and DPD can produce false negative results more frequently than SNA, that is, reporting that nerves are healthy while in reality they already have existing neurological changes, implying that some individuals may not receive the proper treatment as early as possible.

The negative likelihood ratio confirms that the DPD produces more false negatives than SNA, so when the DPD tells us that there are no neurological changes yet, they may already be present at a more subtle level, so we suggest that the DPD always be performed after the results of the SNA and not determining the DPD using only the table that helps in the summary of the SNA data proposed by the guidelines, which considers visible deficiencies such as a Degree of Physical Disability Level 2 (DPD 2) or a barely visible Degree of Physical Disability level 1 (DPD 1), since neurological alterations are thus underestimated.

The WHO grades has been used as an indicator for early case detection and as an indicator for change in impairments, but depending on the expertise of the team, the degree of disability will be the only instrument used to monitor and assess the patient, and The WHO Grades are not designed to be sensitive to early changes.

Greater coverage is achieved, for the early identification of neural injuries, by applying the 3 assessments studied in this research, opting for SNA, if you choose only one, as it was the most sensitive and accurate instrument for identifying neural injuries in isolation, in addition to having the least negative likelihood ratio.

NDA does not establish a relationship of dependency with DPD instruments, nor with SNA; in its application to investigate neural injury in people who have leprosy, it does not prove to be such a sensitive tool in isolation, but when associated with clinical anamnesis and evaluations already used, DPD and SNA, it facilitates diagnosis, impacting the suspicion of new cases of the disease.

The limitation of this study is found in the patient’s lack of access to more complex tests such as electroneuromyography or ultrasound. The fact that the treatment is still centralized, makes it difficult for the patient to adhere, especially those who have subtle neurological changes that already suggest neural damage, but the patient will hardly seek the health system if he does not notice the symptoms.

The association of leprosy and neurodynamics issues alerts professionals not involved with leprosy to suspect leprosy neuropathy when they find a positive neurodynamic test in their clinical practice. Neurodynamic tests were positive in 7.4% of individuals while there were still no changes in SNA and later these changes appeared. Future studies could clarify whether NDA can reveal neural injury earlier.

## Data Availability

Data cannot be shared publicly because
of our institution does not provide a repository. Data are available from the authors.

## APPENDIX 1

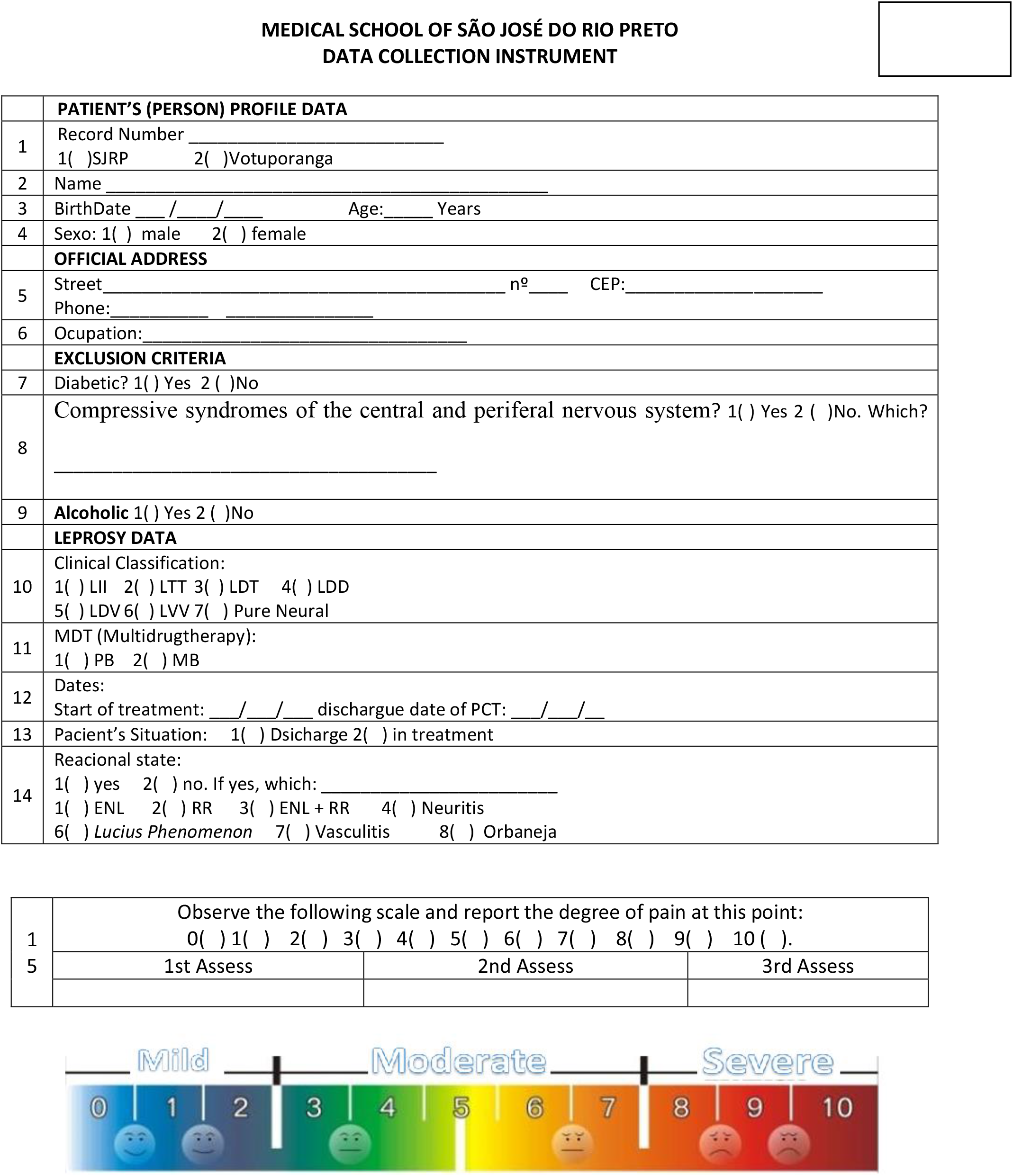
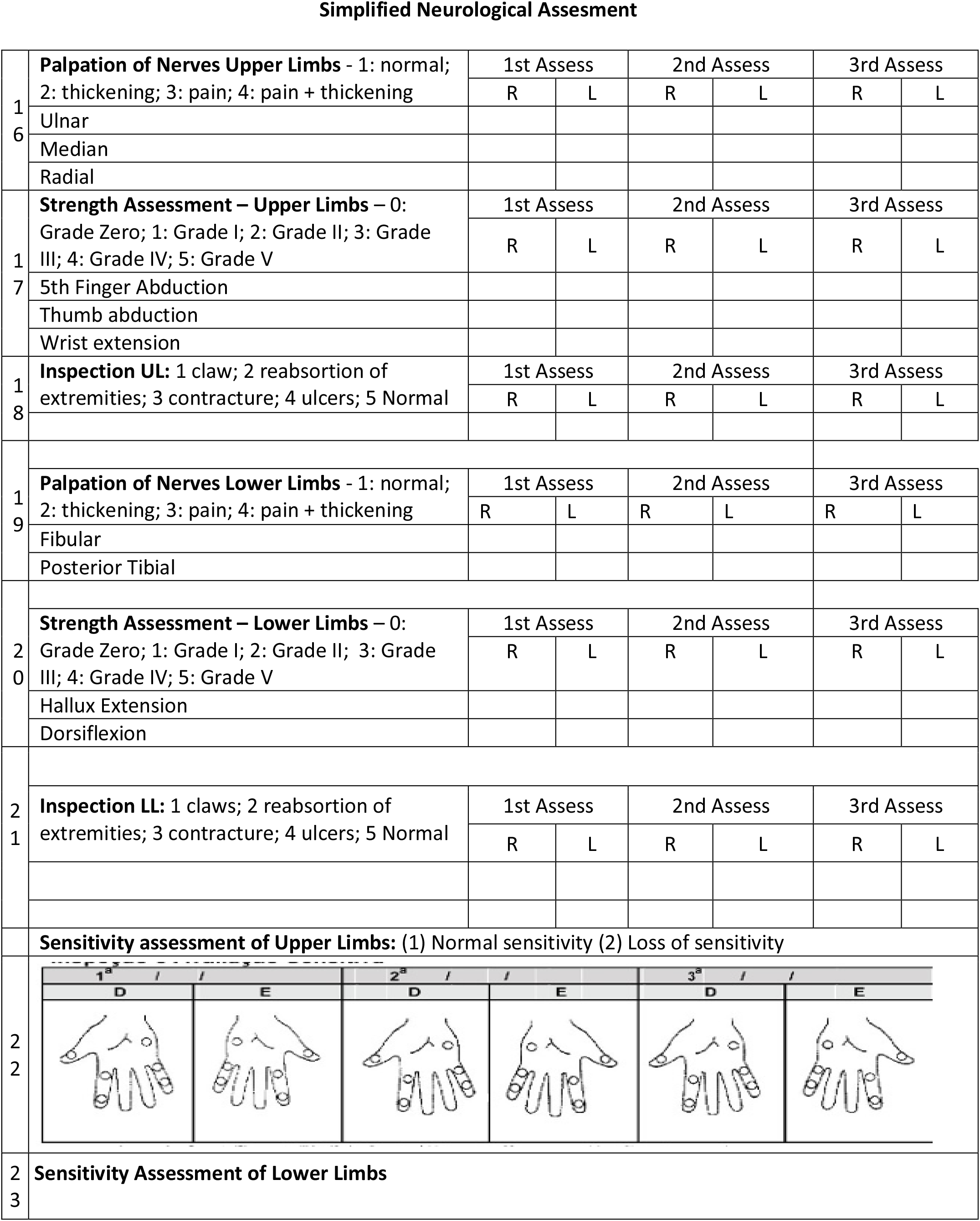
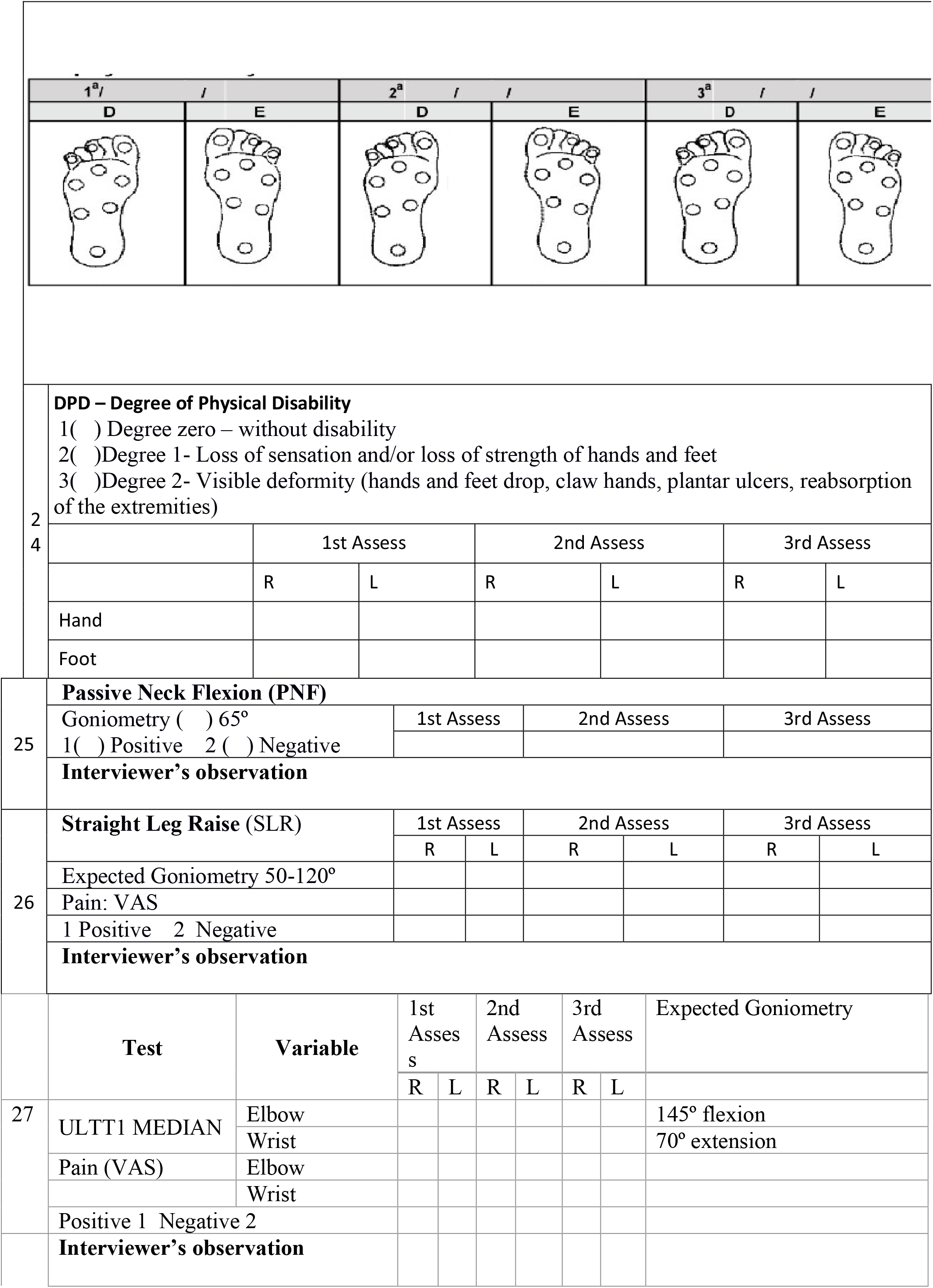
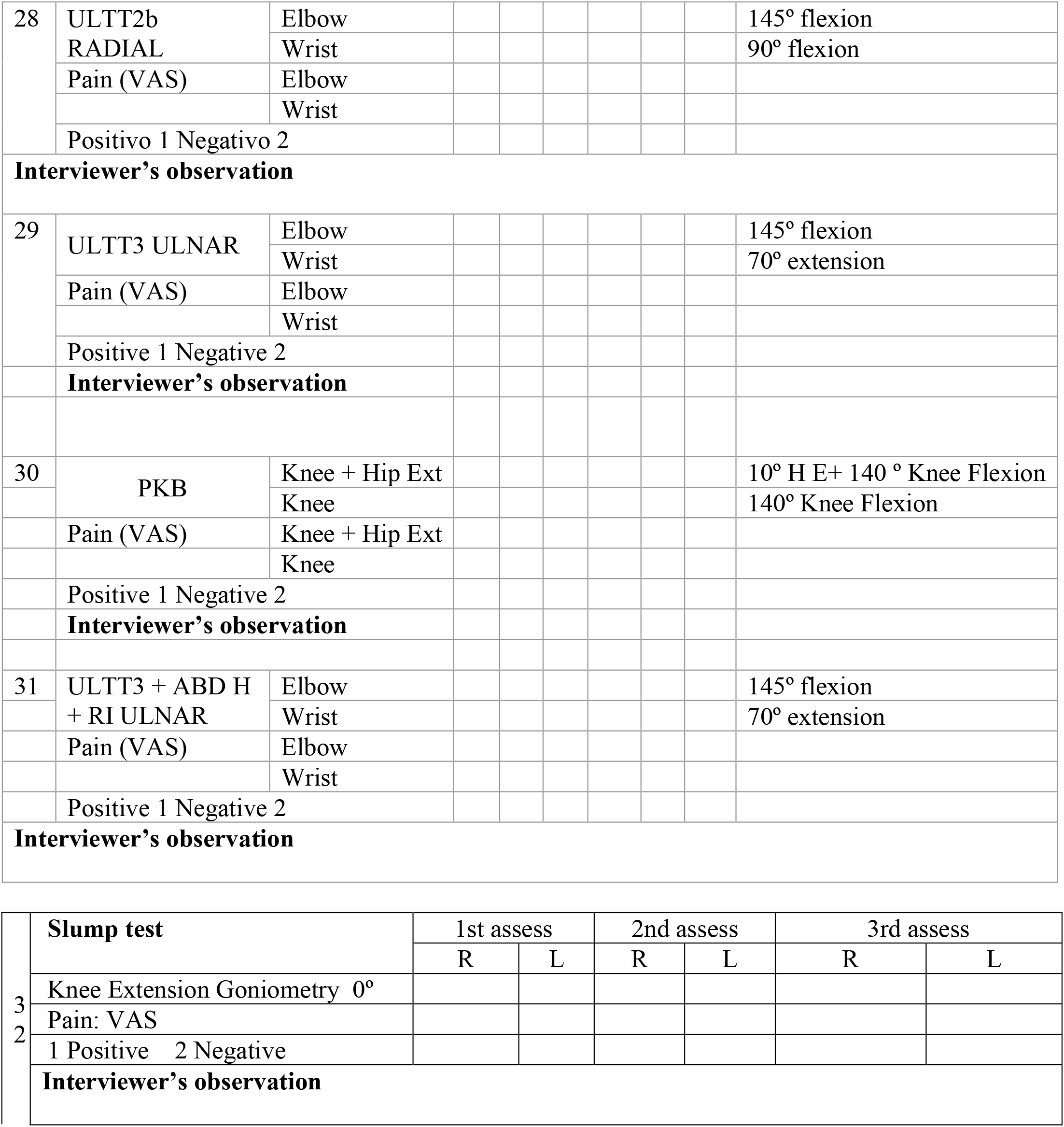

